# ANALYSIS OF RIFAMPICIN INDETERMINATE RESULTS USING SHEWHART CONTROL CHARTS: IMPLICATIONS FOR PATIENTS AND TUBERCULOSIS CONTROL PROGRAMMES

**DOI:** 10.1101/2023.02.21.22282624

**Authors:** Taagbara Jolly Abaate, Aloni Adolphus Alali

## Abstract

**Background:** Antimicrobial resistance is a growing global public health concern, and multidrug-resistant tuberculosis is responsible for roughly one-quarter of all antimicrobial-resistant infection-related deaths worldwide. GeneXpert is a rapid, automated molecular test that detects multi-drug-resistant tuberculosis using rifampicin as a predictor. It was recommended by the World Health Organization (WHO) in 2010 for national tuberculosis programs in developing countries; however, it has limitations. Indeterminate results for Mycobacterium tuberculosis indicate that the test was unable to determine whether the bacteria were resistant to rifampicin. This study used Shewhart Control Chart, which has action limits, to investigate the causes of indeterminate results.

**Methods:** The control limits on the Shewhart chart are central, upper, and lower. GeneXpert indeterminate results obtained between January 2017 and December 2020 in a tertiary hospital in a low and middle-income country were plotted. Points above the upper control limit were used to determine whether or not the process was under control.

**Result:** The proportion of GeneXpert results that were indeterminate varied, with 58% exceeding the upper limit. Only 42% were within the control limit, in comparison.

The majority of the laboratory results revealed an out-of-control signal by displaying points outside the control limits or non-random patterns of points known as special-cause variation, according to this study.

**Conclusions:** GeneXpert indeterminate results have an impact on patient management by preventing accurate diagnosis and delaying the start of anti-tuberculosis medication. Machine malfunctions, insufficient bacterial load, poor quality samples, operator errors, or faulty laboratory materials could all be to blame. Regular equipment checks by laboratory personnel, program sponsors, or leadership will be extremely beneficial in achieving the desired results and initiating appropriate treatment. Statistical process control is widely used in hospital performance monitoring and improvement, and it is becoming more popular in public health surveillance.

## BACKGROUND

With an estimated 10.4 million TB cases and 600,000 new drug resistance cases worldwide1-7, tuberculosis (TB) kills more people than any other single infection. The global burden of tuberculosis cases and drug resistance is increasing 1, 2, and many patients continue to rely solely on an ineffective diagnostic test developed more than a century ago3.

Tuberculosis is still a major public health concern around the world, with over 90% of TB cases occurring in low and middle-income countries like Nigeria.

4 In 2018, the 30 most epidemiologically burdened TB countries accounted for 87% of new TB cases, with eight of these countries, including Nigeria, accounting for two-thirds of new TB cases. 6 Nigeria still has the highest TB burden in West Africa. 4, 5 it is also one of 14 countries with a high burden of TB,

TB/HIV, and Multi-Drug Resistant TB. 2, 6 Rapid global increases in tuberculosis (TB) and multidrug-resistant tuberculosis (MDR-TB) have highlighted the importance of rapid diagnostic techniques. Recent molecular diagnostic techniques have been recommended due to their quick turnaround time and high sensitivity and specificity.8

A molecular diagnostic technique; GeneXpert MTB/RIF (Cepheid Inc.) is a rapid, automated test for MTB detection and RIF (rifampicin resistance) 8-10. The assay is based on nested real-time PCR and molecular beacon technology. In less than two hours, the results are obtained10. The World Health

The organization (WHO) recommended the GeneXpert MTB/RIF assay in 2010 and continues to do so for national tuberculosis programs in developing countries.9 The gene Xpert MTB/RIF assay can detect potential MDR-TB immediately, breaking the barrier of delayed diagnosis in MDR-TB control. 3, 5

Because RIF resistance usually coexists with INH resistance, it is a predictor of MDR-TB. 9, 10 Rapid RIF resistance diagnosis allows TB patients to begin effective treatment sooner than other types of drug susceptibility testing6. Rapid results from the gene Xpert MTB/RIF assay may help patients who do not have tuberculosis save money by avoiding unnecessary treatment and respiratory isolation in hospitals or other institutions8. The results of the gene Xpert MTB/RIF assay indicate whether MTB was detected in the sample, and in some cases, the result is “invalid,” indicating that the test should be repeated. If MTB was discovered, the results will show whether resistance to RIF was discovered, not discovered, or was indeterminate3-9.

Positive GeneXpert results for MTB with RIF resistance indicate that the bacteria are likely to be RIF resistant, which should ideally be confirmed by additional rapid sensitivity testing9-12. However, because the rapid test is not readily available in Nigeria, most Directly Observed Treatment Short Course (DOTS) centers refer patients to MDRTB centers when RIF resistance is detected using GeneXpert molecular testing. 13-17 Positive results for MTB but indeterminate for RIF resistance indicate that the test was unable to accurately determine whether the bacteria were resistant to RIF; therefore, in such cases, growth-based susceptibility testing to first-line TB drugs should be performed, 15-18.

Although the gene Xpert MTB/RIF test represents a significant advancement in TB diagnostic testing, it does have limitations, including a limited shelf-life for diagnostic cartridges, some operating temperature and humidity restrictions, the need for electricity, unknown long-term robustness, and the requirement for annual servicing and calibration of each machine. 3-14. Laboratories in low and middle-income countries, such as Nigeria, are littered with expensive equipment that is no longer functional because it was donated to the incorrect setting. Ensuring the long-term availability of servicing and consumables may be more important and difficult than implementing the diagnostic equipment itself. 16, 19

Rifampicin resistance detection, as well as early diagnosis and improved case detection, are critical to improving individual patients’ health and preventing MDR-TB transmission in the community.16

Tuberculosis is diagnosed using smear microscopy in 24-28 hours, whereas solid culture takes 4-8 weeks.

Early diagnosis and treatment are hampered by this diagnostic lag.

Similarly, test failures or unsuccessful test results caused by process errors delay or prevent some patients from receiving a diagnosis. During the early stages of geneXpert15-17 implementation, other countries expressed similar concerns about high rates of error and invalid results.

In the laboratory, temperature-related, technical problems, cartridge malfunction, electrical connection, and machine malfunction errors are all possibilities18. Other sources of inaccurate Xpert results have been identified as human errors such as noncompliance with Standard Operating Procedures (SOP) during sample processing, filling reaction tubes with viscous sputum or incorrect sample volumes, clogged filters due to the presence of debris in sputum, compromised probe integrity, and module failures18-20. 19-21

According to one study, 20 high rates of failed test results demonstrated that early diagnosis, which is critical to TB control, may not be possible in many people if gene Xpert is not managed properly.

Patients who receive false positive RIF-resistant GeneXpert results are more likely to be treated with second-line anti-TB drugs, which are known to be toxic to patients and have significant financial implications for the national TB program. MTB concentrations of 5, 6, 16, and 20 are low or very low in samples with non-conforming RIF susceptibility reports, implying that bacterial load influences the interpretation of RIF resistance and indeterminate strains.9, 10, 21 Previous research has found that an unfixed bacterial load affects RIF susceptibility status 21. The assay’s ability to distinguish between mutated and wild-type sequences in the rpoB gene’s core region may be limited due to the low bacterial load.

When MTB concentrations are very low and resistance cannot be determined, RIF indeterminate results have also been observed19 21. When running the gene Xpert test, the first and last probes with CT values that exceed the predefined limits may be reported as RIF sensitive18-22. As a result, GeneXpert assay parameters should be reviewed, particularly the CT cut-offs for establishing RIF resistance and indeterminate status.30,32 This is done to lessen the negative effects of misdiagnosis on not only the patient but also on the TB program and the community as a whole. 2 Walter Shewhart variation theory categorizes variation as coming from a “common cause” or a “special cause,” and it guides the appropriate action needed to address these variations and provide continuous improvement. 31.

A common cause variation is expected as a result of chance.31, 39 it is present in all processes and has an impact on everyone involved. We must change the underlying process to reduce common cause variation. 31 Special cause variation, on the other hand, is an unusual variation that does not occur by chance but rather as a result of specific circumstances and thus does not affect everyone involved in the process. Special cause variation can produce either exceptionally good or exceptionally bad results31, and to reduce unfavorable special cause variation, we must first identify it and then act on it. We must also investigate and learn from favorable special cause variations for the benefit of others. 29, 31

Control charts have been proposed for use in clinical governance and performance monitoring in healthcare.28, 29. The inconsistency and indeterminacy of the gene Xpert assay, which is commonly seen in the DOTS clinic at a tertiary healthcare facility in Rivers state Nigeria, were investigated using control charts. The goal of this paper is to describe a method for evaluating variation in GeneXpert proportions and to attempt to quantify inconsistencies in laboratory GeneXpert results.

## THE OBJECTIVE OF THE RESEARCH

The purpose of this research was to determine whether the output of the GeneXpert process in DOTS at a tertiary center in Port Harcourt, Rivers state, Nigeria, was under control, and if so, whether the variations (if any) in Rifampicin indeterminate GeneXpert results from these laboratories were due to special course variations or common course variations.

## METHODS

GeneXpert data routinely collected from hospital laboratories in DOTS clinics were obtained. The data included the number of sputa taken per month and the number of indeterminate results observed in the same month from 2017 to 2020. We investigated the internal consistency of the laboratory GeneXpert results using P-charts. P-charts are a type of control chart that is intended to be used with binomial data (yes or no) expressed as a proportion of the sample size. 33-35 we ordered our data by the total number of results and plotted our P-charts accordingly. One advantage is that the resulting P-charts produce visually appealing control limits because they resemble a funnel. Such plots have been proposed in the medical field.30 SPSS version 23.0 was used for all analyses.

## RESULTS

The proportion of indeterminate GeneXpert results varied, with 58% exceeding the upper limit. In comparison, only 42% were within the control limit.

## DISCUSSION

Control chart-based monitoring begins with selecting the type of chart to use. To investigate inconclusive gene Xpert results, the Shewhart control chart, a simple, low-cost, effective tool with high sensitivity and specificity, was used. During the study period, an estimated 58% of total results were outside the upper control limit, while 42% were within it. During the study period, the majority of the laboratory’s results showed an out-of-control signal, indicating a faulty process that could have been caused by machine issues, operator errors, sputum quality, or faulty laboratory materials. On a control chart for a controlled process, points are distributed at random. In other words, it only differs from sources common to the process (called common-cause variation). Figure 1 depicts an out-of-control process by displaying points that are outside the control range or non-random patterns of points (referred to as special-cause variation).

**Figure 1.**
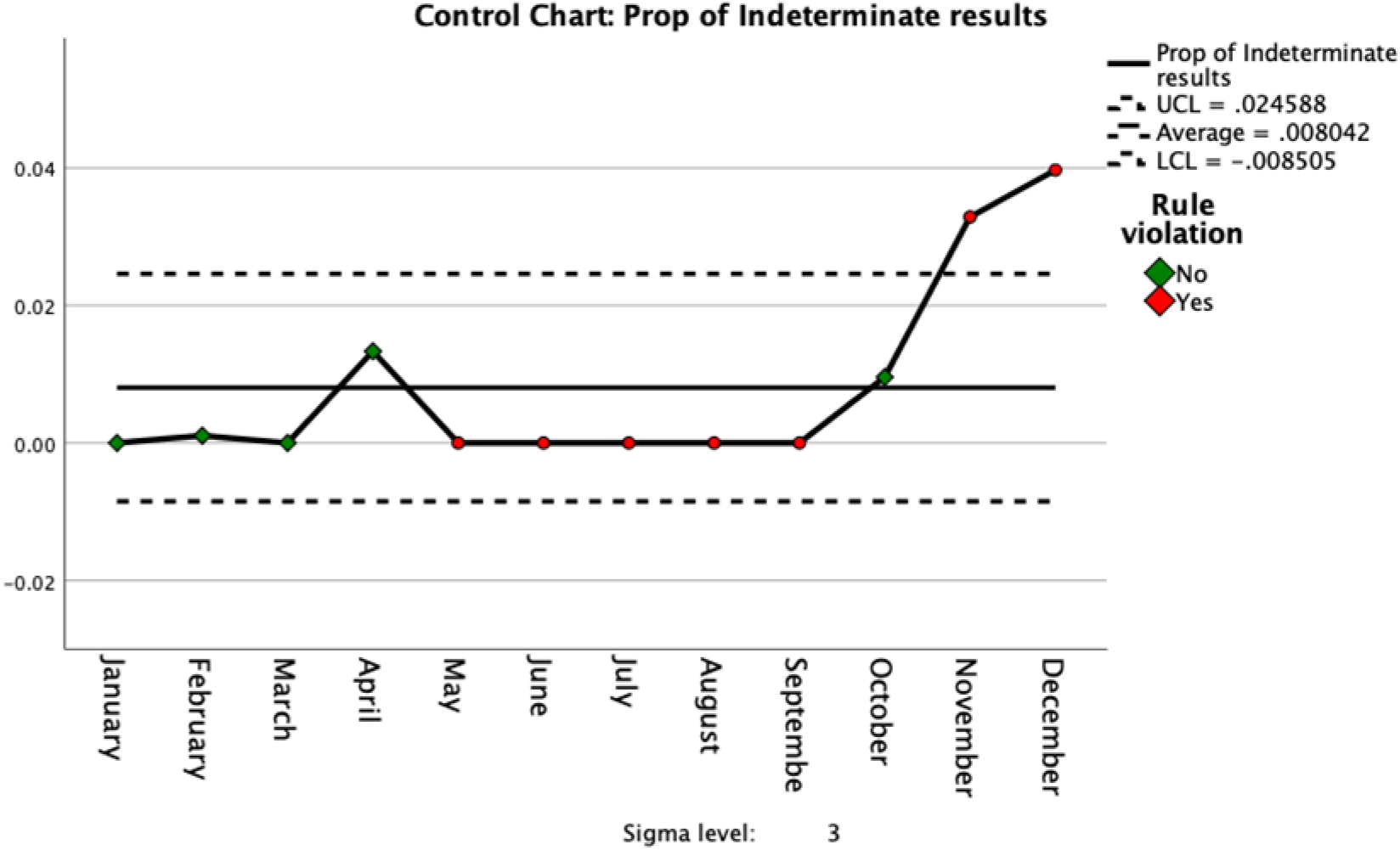
Control chart with five successive points on the same side of the centerline

Patients who could have benefited from early resistance detection and MDRTB treatment may be delayed as a result of an incorrect indeterminate result. As a result, the disease burden will worsen and the program’s costs will rise.

Shewhart control charts were used in a UK study to analyze inadequate cervical smears, and the results guided quality improvement. 31 While similar methodologies for quality control of Xpert results have not been used in Nigeria or elsewhere, statistical process control has been widely used in hospital performance monitoring and improvement and is increasingly being adopted for public health surveillance. 33-35

## STRENGTHS AND LIMITATIONS OF THIS STUDY

The ultimate goal of data is to provide a foundation for action to support improvement, and this study sheds light on the clinical quandary of starting anti-tuberculosis treatment. Shewhart control charts have high sensitivity and specificity, and they are the most common and easiest to build. They work by plotting one data point at a time.

Because this was a population-based study, our findings reflect the quality control issues that most reference laboratories face. Control charts are unpopular among researchers, and this is the first study to use a statistical control tool to examine the stability of quality control measures in the laboratory.

Due to the complexities and time constraints of this study, we did not use the CUSUM charts, which have higher sensitivity and specificity (35, 39).

Because this was a total population study, our findings reflect normal laboratory operations. Points could have strayed beyond the upper or lower control limits, resulting in a false alarm or type 1 error.

The presence of five consecutive points with more than one standard deviation and greater than three sigmas on the same side of the centerline was used to identify special cause variation. Other, more sensitive, and specific detection rules have been proposed. 36, 37, and should be investigated further in future research.

## CONCLUSION

GeneXpert indeterminate results have an impact on patient management by preventing accurate diagnosis and delaying the start of anti-tuberculosis medication. Machine malfunctions, insufficient bacterial load, poor quality samples, operator errors, or faulty laboratory materials could all be to blame. Regular equipment checks by laboratory personnel, program sponsors, or leadership will be extremely beneficial in achieving the desired results and initiating appropriate treatment. Statistical process control is widely used in hospital performance monitoring and improvement, and it is becoming more popular in public health surveillance.

## Data Availability

All data produced in the present study are available upon reasonable request to the authors

## ETHICAL APPROVAL

The Research and Ethics Committee of the University of Port Harcourt Teaching Hospital in Rivers state, Nigeria, granted ethical approval. The authorization ID is UPTH/ADM/90/S.II/VOL.XI/1470.

## DISSEMINATION

The findings from this study will be presented at both local and international conferences and published in key peer-reviewed journals.

## ACKNOWLEDGMENTS

We sincerely thank the staff of the DOTs clinic for collecting the data and making it available for analysis.

## CONTRIBUTORS

ATJ designed and conceptualized the study methodology, while AA performed the primary statistical analysis. All authors approved the final version of the manuscript, and they all contributed significantly to its revision and editing. The corresponding author certifies that all listed authors meet the requirements for authorship and that no other authors who meet the requirements were overlooked.

## PROVENANCE AND PEER REVIEW

Not commissioned; peer-reviewed externally.

